# LONG-TERM PHYSICAL AND MENTAL HEALTH IMPACT OF COVID-19 ON ADULTS IN ENGLAND: FOLLOW UP OF A LARGE RANDOM COMMUNITY SAMPLE

**DOI:** 10.1101/2023.04.24.23289043

**Authors:** Christina J Atchison, Bethan Davies, Emily Cooper, Adam Lound, Matthew Whitaker, Adam Hampshire, Adriana Azor, Christl Donnelly, Marc Chadeau-Hyam, Graham S Cooke, Helen Ward, Paul Elliott

**Affiliations:** School of Public Health, Imperial College London, UK; Imperial College Healthcare NHS Trust, UK; MRC Centre for Environment and Health, School of Public Health, Imperial College London, UK; Department of Brain Sciences, Imperial College London, UK; Department of Statistics, University of Oxford, Oxford, UK; MRC Centre for Global infectious Disease Analysis and Abdul Latif Jameel Institute for Disease and Emergency Analytics, Imperial College London, UK; Health Data Research (HDR) UK London at Imperial College; Department of Infectious Disease, Imperial College London, UK; National Institute for Health Research Imperial Biomedical Research Centre, UK; UK Dementia Research Institute at Imperial College, UK

## Abstract

**Background:** The COVID-19 pandemic is having a lasting impact on health and well-being. We compare current self-reported health, quality of life and symptom profiles for people with ongoing symptoms following COVID-19 to those who have never had COVID-19 or have recovered.

**Methods:** A cohort study was established with participants from the REACT programme. A sample (N=800,000) of adults were contacted between August and December 2022 to complete a questionnaire about their current health and COVID-19 history. We used logistic regression to identify predictors of persistent symptoms lasting ≥12 weeks following COVID-19. We fitted Accelerated Failure Time models to assess factors associated with rate of recovery from persistent symptoms.

**Findings:** Overall, 276,840/800,000 (34.6%) of invited participants completed the questionnaire. Median duration of COVID-related symptoms (N=130,251) was 1.3 weeks (inter-quartile range 6 days to 2 weeks), with 7.5% and 5.2% reporting ongoing symptoms ≥12 weeks and ≥52 weeks respectively. Female sex, having ≥1 comorbidity, more severe symptoms at time of COVID-19 and being infected when Wild-type variant was dominant were associated with higher probability of symptoms lasting ≥12 weeks. Longer time to recovery in those with persistent symptoms was found for females, people with comorbidities, living in more deprived areas, current smokers and for Wild-type compared to later variants. Mental health and health-related quality of life were significantly worse among participants with ongoing persistent COVID-19 symptoms compared with those who had never had COVID-19 or had recovered.

**Interpretation:** Although COVID-19 is usually of short duration, some adults experience persistent and burdensome illness.

**Funding:** This work is independent research funded by the National Institute for Health and Care Research (NIHR) (REACT Long COVID (REACT-LC) (COV-LT-0040)). This research is part of the Data and Connectivity National Core Study, led by Health Data Research UK in partnership with the Office for National Statistics and funded by UK Research and Innovation (UKRI) (MC_PC_20029). The views expressed in this publication are those of the author(s) and not necessarily those of NIHR or UKRI.

## INTRODUCTION

The UK has experienced one of the largest epidemics of COVID-19 (symptomatic SARS-CoV-2 infection) in Europe. As well as the risk of hospitalisation and death from COVID-19 it is clear that some people who develop symptoms have a prolonged and debilitating illness that may continue for weeks or months (Long COVID or post-COVID syndrome) (1, 2). Pathogenesis of persistent symptoms is poorly understood and represents a major knowledge gap if effective treatments and management strategies are to be developed.

Our initial estimates from the REal-time Assessment of Community Transmission-2 (REACT-2) study, six rounds of repeat cross-sectional random samples of the population to evaluate community prevalence of SARS-CoV-2 anti-spike protein antibody positivity in England (3), suggested that 21.6% of adults with evidence of prior infection experience one or more symptoms 12 weeks after their initial illness (4). With 3.4 million individuals thought to have been infected during the first peak in England alone (5), and significant transmission since, there may be long-term major challenges to healthcare services even with rapid scale up of effective vaccination. Long COVID may require new treatment approaches, and better diagnostic and prognostic signatures will be vital for more effective management.

The REACT programme is one of the world’s largest and most comprehensive coronavirus monitoring studies. In addition to REACT-2 described above, the REACT-1 study included 19 rounds of repeat cross-sectional random samples of the population to track community SARS-CoV-2 infection with PCR tests (3). The REACT programme is uniquely placed to identify individuals with persistent symptoms who have not been hospitalised and to compare them with people whose symptoms have resolved and those who have never had COVID-19.

Here, we use follow up data from a survey of REACT participants to assess the impact of the pandemic on the health and wellbeing of people in England between August and December 2022, and describe how this varies according to their experience of COVID-19. We report the duration of symptoms in people with a history of symptomatic infection, assess factors associated with symptom persistence beyond 12 weeks (Long COVID) and with recovery after that point. We also compare current self-reported health and quality of life and specific symptoms for those with Long COVID to those who have never had COVID-19 or have recovered.

## METHODS

The REACT programme sampled random cross-sections of the population in England to quantify community prevalence of virus by RT-PCR (REACT-1, 19 rounds between May 2020 and March 2022) and of IgG anti-SARS-CoV-2 antibody based on a self-administered lateral flow immunoassay (LFIA) test (REACT-2, six rounds between June 2020 and May 2021) (3). Methods for the studies are published elsewhere (3, 6). In addition, participants provided information on demographics, household composition, comorbidities, and COVID-19 history. Those who reported having had COVID-19 were asked whether they had had a PCR test, symptoms related to COVID-19, date of first symptom onset, severity of symptoms, and duration of any of a list of 29 symptoms. The questionnaires used and a table showing the sampling dates for each study round, and the response rates are available on the study website (7). The average response rate across both studies and all rounds was 23.4%. Each round of data collection included between 90,000 and 210,000 participants. Overall, 3,099,386 adults registered to take part in the REACT programme. Of these participants, 80.5% (2,494,309/3,099,386) consented to both recontact and data linkage of routine health records.

### Study design and participants

In this follow-up study (see supplementary figure S1) we aimed for a sample size of at least 160,000; assuming a 20.0% response rate, we obtained a sample of 800,000 adults aged ≥18 years using as a sample frame all REACT-1 and REACT-2 participants who had consented to both re-contact and data linkage (n= 2,494,309). Personalised invitations were sent via email. To increase our sample of individuals with persistent symptoms of COVID-19 we first invited all individuals in the following subgroups:

1. Individuals from REACT-1 or REACT-2 with a previous history of self-reported test confirmed or suspected COVID-19 who reported persistent symptoms of ≥12 weeks (n=52,501)
2. Individuals from REACT-1 who tested positive for SARS-CoV-2 as part of the study (n=13,482)
3. Individuals from REACT-2 who tested positive for SARS-CoV-2 IgG as part of the study and had not been vaccinated at the time (n=85,757)

To achieve the 800,000-participant size, a random sample (n=648,260) of all remaining adults not meeting the above criteria was selected.

Participants registered via an online portal. Those registered completed an online questionnaire (7). The questionnaire was designed, piloted and revised with input from our public advisory group including people with Long COVID. It was designed to collect data on current health, well-being, functionality and recent symptoms, followed by questions about history of COVID-19 (SARS-CoV-2 PCR and lateral flow devices (LFD) test results, frequency, severity, duration). This was to compare current health status, including symptoms, which were assessed before people were asked about possible persistence of COVID-19 symptoms. Current health status included a set of 29 symptoms potentially related to COVID-19 including: (i) loss or change of sense of smell and taste, (ii) coryzal symptoms, (iii) gastrointestinal symptoms, (iv) fatigue-related symptoms, (v) respiratory and cardiac symptoms (vi) memory and cognitive symptoms, (vii) other flu-like and miscellaneous symptoms. In addition to questions regarding fatigue and sleep quality, mental and physical health outcomes were collected using the following validated questionnaires:

1. Quality of life/functioning: EuroQol five-dimension five-level (EQ-5D-5L) (8)
2. Assessment of breathlessness in people reporting this symptom: Dyspnoea-12 (9)
3. Assessment of post-exertional malaise (PEM) in people reporting fatigue: three PEM-items from the DePaul Symptom Questionnaire (10)
4. Mental health: Generalized Anxiety Disorder 7-item scale (GAD-7) (11) and Patient Health Questionnaire-9 (PHQ-9) (12)

On completion of the questionnaire, participants were invited to complete a 15-minute online cognitive assessment (13).

### Data linkage

The UK Health Security Agency (UKHSA) receives results of all SARS-CoV-2 PCR tests in England from community settings (Pillar 2) (14). In addition, members of the public are encouraged to submit results of at-home self-testing using LFDs (Pillar 2) (14). To obtain additional information on dates of positive SARS-CoV-2 tests, participant study data were linked to their Pillar 2 records using their unique National Health Service (NHS) number and other personal identifiers.

To obtain information on dates of received COVID-19 vaccine doses, participant study data were linked to their NHS records from NHS Digital on COVID-19 vaccination events in any setting (15). This was done using their unique NHS number and other personal identifiers.

### Data Analysis

Our primary analyses focused on prevalence of individual symptoms currently reported at the time of questionnaire completion and validated self-reported physical and mental health outcome measures by the following COVID-19 categories of participants:

1. **No COVID**: no history or evidence of SARS-CoV-2 infection or COVID-19;
2. **Asymptomatic or resolved short COVID <4**: Asymptomatic SARS-CoV-2 infection or COVID-19 symptoms resolved within 4 weeks;
3. **Resolved short COVID** ≥**4 to <12**: COVID-19 symptoms resolved within 4-12 weeks;
4. **Resolved persistent COVID**: COVID-19 symptoms lasting ≥12 weeks but no longer symptomatic; this was further divided into those lasting less than 52 weeks and those lasting ≥52 weeks before resolution.
5. **Ongoing persistent COVID**: COVID-19 symptoms lasting ≥12 weeks and ongoing; this was further divided into those lasting less than 52 weeks to date and those lasting ≥52 weeks to date.

We included only symptomatic SARS-CoV-2 infections confirmed by a positive test result (PCR or LFD) in our definition of COVID-19 for our main analyses. This included self-reported test positives (survey question), REACT-1 test positives, unvaccinated REACT-2 SARS-CoV-2 IgG test positives and Pillar 2 test positives. Asymptomatic infections were defined as test (PCR or LFD) positives with no reported symptoms. A repeat positive test result was included as a separate infection if performed ≥90 days after a previous positive test (16). The COVID-19 episode date used was symptom onset date for symptomatic infections or date of a positive SARS-CoV-2 PCR or LFD test for asymptomatic infections. We excluded individuals with less than 12 weeks follow up from their COVID-19 episode date to their questionnaire completion date.

Index of Multiple Deprivation (IMD) 2019 was used as a measure of relative deprivation, based on seven domains at a community level (Lower Layer Super Output Area, approximately 1,500 residents) across England (income, employment, education, health, crime, barriers to housing and services, and living environment) (17). Participants were allocated to quintiles of deprivation based on their residential postcode. A valid COVID-19 vaccine dose was defined as a date of vaccination 14 days or more prior to the COVID-19 episode date. The Wild-type strain was dominant in the UK prior to December 2020. Alpha dominated between December 2020 and April 2021 followed by Delta (May 2021 to mid-December 2021) and Omicron (late December 2021 onwards) (18).

Continuous variables were presented as median (IQR) or mean (SD), as appropriate. Binary and categorical variables were presented as counts and percentages. A χ² test was used to identify differences in proportions across COVID-19 categories. For normally distributed continuous data, analysis of variance (ANOVA F-test) was used to test differences across categories, with Kruskal-Wallis tests used for non-normally distributed data.

We used logistic regression (adjusted Odds Ratios (aOR) and 95% Confidence Intervals (CI) adjusted for age, sex, ethnicity, IMD, comorbidities (presence of a pre-existing health condition) and smoking status) to compare current self-reported specific symptoms for those with ongoing persistent COVID-19 symptoms to those who had never had COVID-19 or had recovered. Further, we used mutually logistic regression to quantify the associations of age, sex, ethnicity, IMD, comorbidities, smoking status, severity of initial illness, COVID-19 vaccination status, and dominant UK circulating SARS-CoV-2 variant at time of infection with COVID-19 symptoms lasting ≥12 weeks and lasting ≥52 weeks.

The dataset was converted into a format suitable for survival analysis techniques. Participants were followed up from their COVID-19 symptom onset date until the reported symptom end date (participants provided one date for when all symptoms had resolved) or, if they reported ongoing symptoms, until the survey completion date. We constructed Kaplan-Meier plots of time to symptom end date. To assess factors associated with symptom recovery in participants with symptom persistence beyond 12 weeks, we used an Accelerated Failure Time model (19) to quantify the associations between COVID-19 symptom discontinuation beyond 12 weeks and the following factors: age, sex, ethnicity, comorbidities, IMD, smoking status, severity of acute illness, COVID-19 vaccination status, and dominant UK circulating SARS-CoV-2 variant at time of symptom onset. Mutually adjusted Time Ratios (aTR) and 95% Confidence Intervals (CI) were estimated. A TR >1 is interpreted as a slower symptom recovery rate beyond 12 weeks in participants with COVID-19 symptoms lasting ≥12 weeks. For example, a “Female” TR of 1.10 where Males are the baseline group is interpreted as symptoms resolved at a 10% slower rate in females compared to males.

All tests were two-tailed and p values of less than 0.05 were considered statistically significant. We did not adjust for multiple testing. Data were analysed using the statistical package STATA version 15.0.

### Ethical approval

Research ethics approval was obtained from the South Central-Berkshire B Research Ethics Committee (IRAS IDs: 298404, 259978, 283787 and 298724).

### Patient and public involvement

A public advisory group provides regular review of the study design, processes and results including these findings and made helpful suggestions for areas of focus including exploring factors associated with recovery.

## RESULTS

Of the 800,000 people who were sent invitations between 1 August–1 December 2022, 282,780 (35.3%) registered for the study, of whom 276,840 (97.9%) completed the questionnaire. Differential non-response was observed by sociodemographic characteristics, including age, sex, ethnicity and deprivation (see supplementary table S1) and were similar to those of the REACT-1 and 2 studies overall (20, 21).

Of those who completed the questionnaire, 266,854/276,840 (96.4%) reported whether they had a confirmed SARS-CoV-2 infection. In total, 59.1% (157,668/266,854) of participants had tested positive for SARS-CoV-2, 19,074/266,854 (7.1%) had two or more test confirmed infections. Overall, 24,142 respondents were excluded because COVID-19 episode date was within 12 weeks of their survey completion date (see supplementary figure S1). Supplementary Table S2 shows the key sociodemographic and COVID-19 categories of participants included in the study.

### Factors associated with persistent COVID-19 symptoms and recovery

There were 130,251 people who reported having had at least one episode of test-confirmed, symptomatic COVID-19. They reported a median duration of COVID-related symptoms of 1.3 weeks, (mean 5.4 weeks, range 1 day to 3.0 years, IQR 6 days to 2 weeks), with 10.2%, 7.5%, and 5.2% reporting ongoing symptoms beyond 4 weeks, 12 weeks and 52 weeks, respectively (Figure 1). Figure 2 shows factors associated with having COVID-19 symptoms persisting for ≥12 weeks (“Long COVID”, LC) and ≥52 weeks (“very Long COVID”, VLC) compared to those who were asymptomatic or whose symptoms resolved within 4 weeks (see supplementary table S3). In these mutually adjusted models, LC and VLC were both associated with being female compared to male (aOR1.42 [95% CI 1.35, 1.50] and 1.49 [1.38, 1.62] for LC and VLC respectively), having ≥1 comorbidities compared to none (aOR 1.31 [1.19, 1.44], 1.52 [1.31, 1.76] for 1, and 1.46 [1.27, 1.75], 2.35 [1.85, 2.97] for ≥2 compared to no comorbidities), and having had moderate or severe symptoms compared to mild at the time of infection (aOR 1.76 [1.63, 1.89], 1.47 [1.32, 1.64] for moderate and 4.87 [4.52, 5.25], 3.55 [3.19, 3.96] for severe). The odds of LC and VLC were lower in people of Asian than white ethnicity (aOR 0.80 [0.69, 0.93], 0.71 [0.57, 0.88]), and in people infected at a time when Alpha (aOR 0.60 [0.56, 0.64], 0.59 [0.54, 0.64]), Delta (OR 0.38 [0.35, 0.41], 0.32 [0.29, 0.36]) and Omicron (OR 0.12 [0.11, 0.13] for LC, insufficient follow-up time for VLC) were dominant compared to Wild-type. There was also a gradient of reducing odds with lower deprivation. LC, but not VLC, was slightly lower in older than younger people.

**Figure 1.**
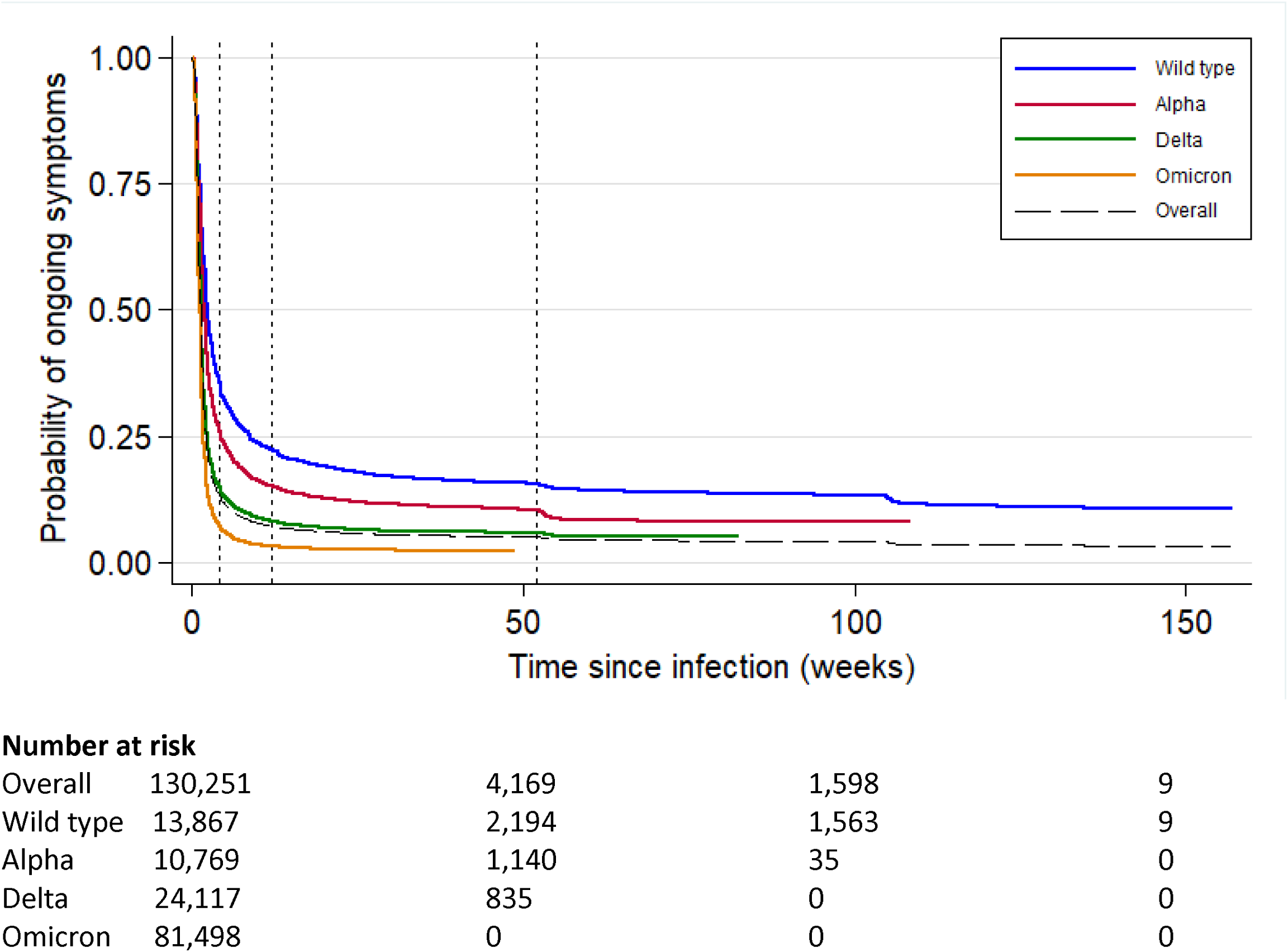
Kaplan Meier Survival Curve of time to symptom end date in those self-reporting symptomatic SARS-CoV-2 infection, overall and by dominant variant at the time of infection.

**Figure 2.**
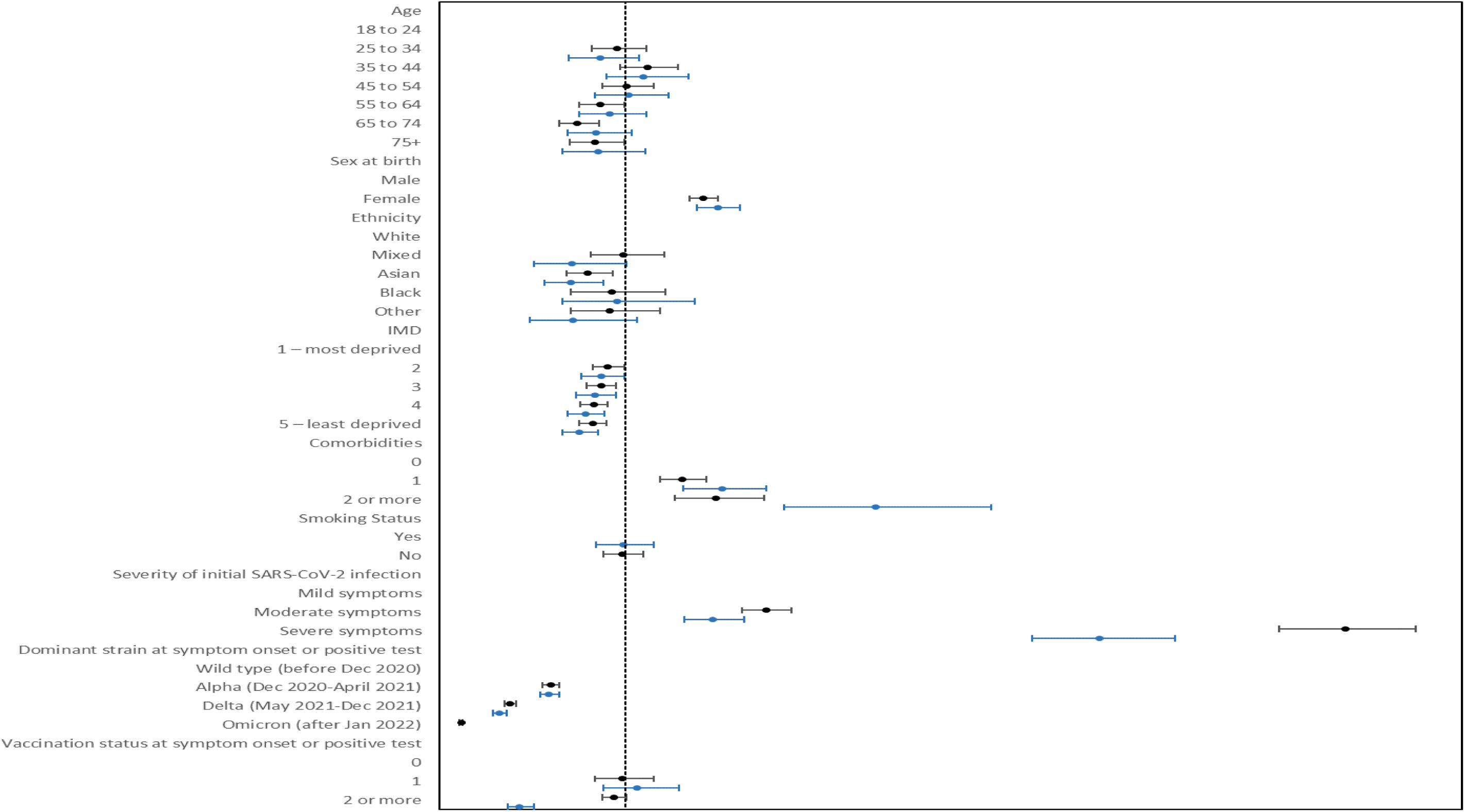
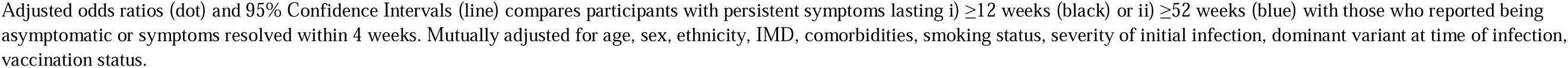
Factors associated with persistent COVID-19 symptoms lasting i) ≥12 weeks and ii) ≥52 weeks versus those who reported being asymptomatic or symptoms resolved within 4 weeks.

For people who had LC (≥12 weeks), we assessed the factors associated with longer symptom duration in an accelerated time failure model, which is a measure of non-recovery (see supplementary table S4). Longer time to recovery was found for females (aTR 1.14 [1.06, 1.23] compared to males) and people with comorbidities (aTR 1.24 [1.08, 1.42] and 2.05 [1.58, 2.66] for 1 and ≥2 compared to none, respectively). Shorter time to recovery of LC was found in people of other and mixed ethnicity (aTR 0.63 [0.45, 0.89] and 0.75 [0.57, 0.99] for other and mixed compared to white), people living in least deprived areas (aTR 0.78 [0.69, 0.88] compared to most deprived), non-smokers (aTR 0.73 [0.62, 0.86] compared to current smokers), later variants at the time of infection (aTR 0.79 [0.72, 0.86], 0.89 [0.79,0.99], 0.69 [0.61,0.78] for Alpha, Delta and Omicron respectively compared to Wild-type).

### Current symptom profile and health-related quality of life

Figure 3 shows the current symptom profile of people who reported ongoing persistent symptoms from their SARS-CoV-2 infection and for those who had recovered or had never had COVID-19. The most common symptoms experienced by individuals with ongoing persistent symptoms were mild fatigue (66.9%), difficulty thinking or concentrating (54.9%) and joint pains (54.6%). However, mild fatigue and joint pains were also common in those with no history of COVID-19 and in those whose COVID-19 symptoms had resolved. The greatest difference in symptom prevalence between those with ongoing persistent symptoms and other participants were for loss or change of sense of smell (aOR 9.31, 95% CI: 8.64, 10.04) and taste (aOR 8.47; 95% CI: 7.85, 9.15), shortness of breath (aOR 6.69; 95% CI: 6.29, 7.12), severe fatigue (aOR 6.19; 95% CI: 5.66, 6.77), difficulty thinking or concentrating (aOR 4.97; 95% CI: 4.68, 5.27), chest tightness or pain (aOR 4.71; 95% CI: 4.37, 5.08) and poor memory (aOR 4.40; 95% CI: 4.15, 4.66) (Figure 3).

**Figure 3.**
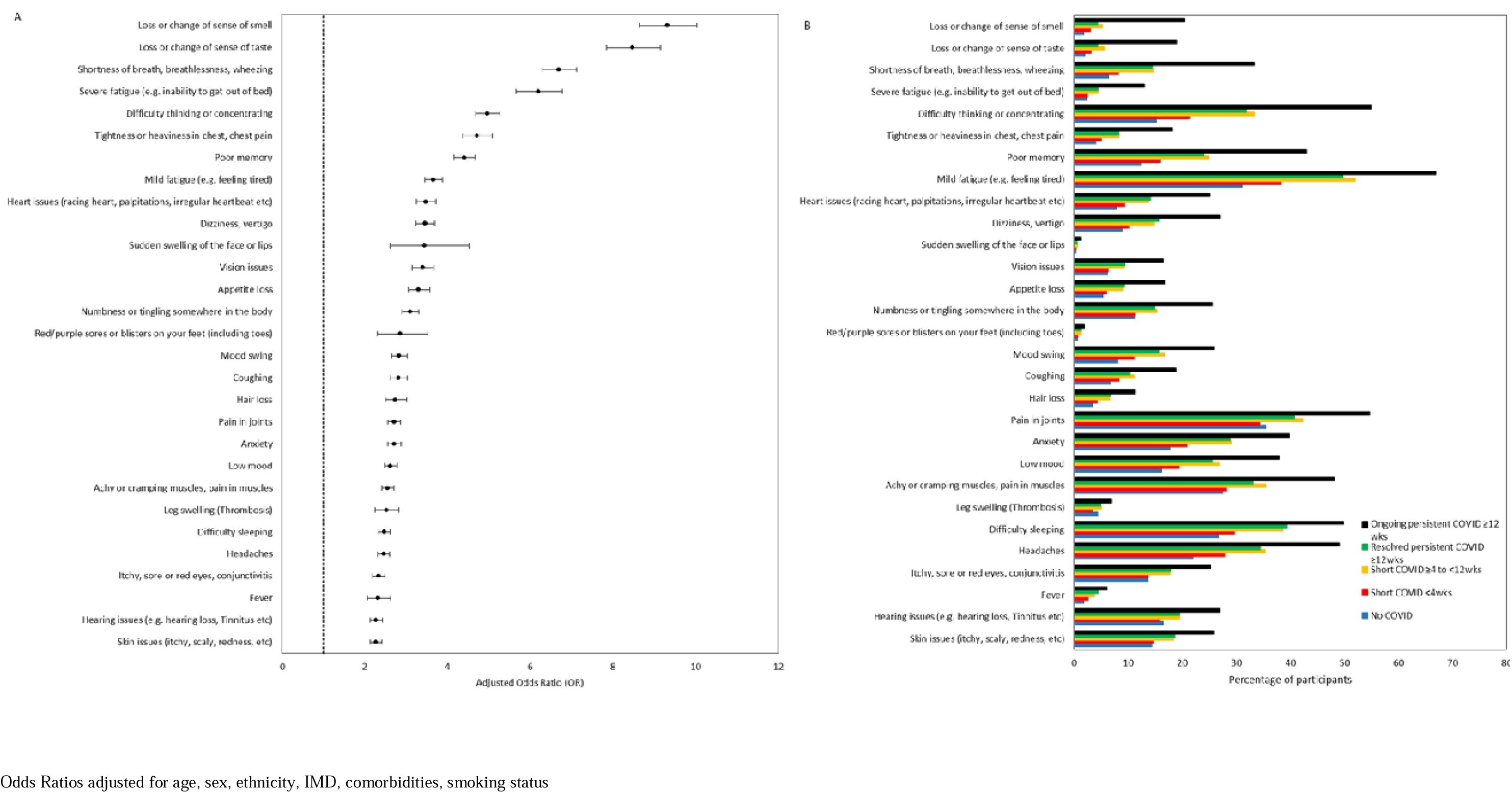
Current symptoms profile by COVID-19 history A) Forest plot of current symptoms in those reporting ongoing persistent COVID-19 symptoms versus all other respondents, and B) Prevalence of current symptoms by COVID-19 history.

Table 1 shows the current symptom profile and health-related quality of life scores of participants by their COVID-19 history. Participants with ongoing COVID-19 symptoms lasting ≥12 weeks reported worse current health, including a higher number of symptoms within the last two weeks, and greater reduction in ability to carry out daily activities due to these symptoms. Post exertional malaise, characterised by asking respondents who reported fatigue about worsening of fatigue symptoms after minimal physical and mental effort, and whether exercise makes fatigue symptoms worse, was also more prevalent in individuals reporting ongoing persistent symptoms post-COVID-19 (Table 1, supplementary table S5). Worse mental health (assessed by PHQ9 and GAD7 scores) and health-related quality of life (assessed by EQ-5D-5L) were reported by participants with ongoing COVID-19 symptoms lasting ≥12 weeks, including problems with mobility, doing usual activities and pain and discomfort (Table 1, supplementary table S5). Table 1 also shows that for the 3,221 people who previously had COVID-19 symptoms lasting ≥12 weeks and report having recovered; their health status is broadly similar to those with shorter recovery or who never had COVID.

**Table 1.**
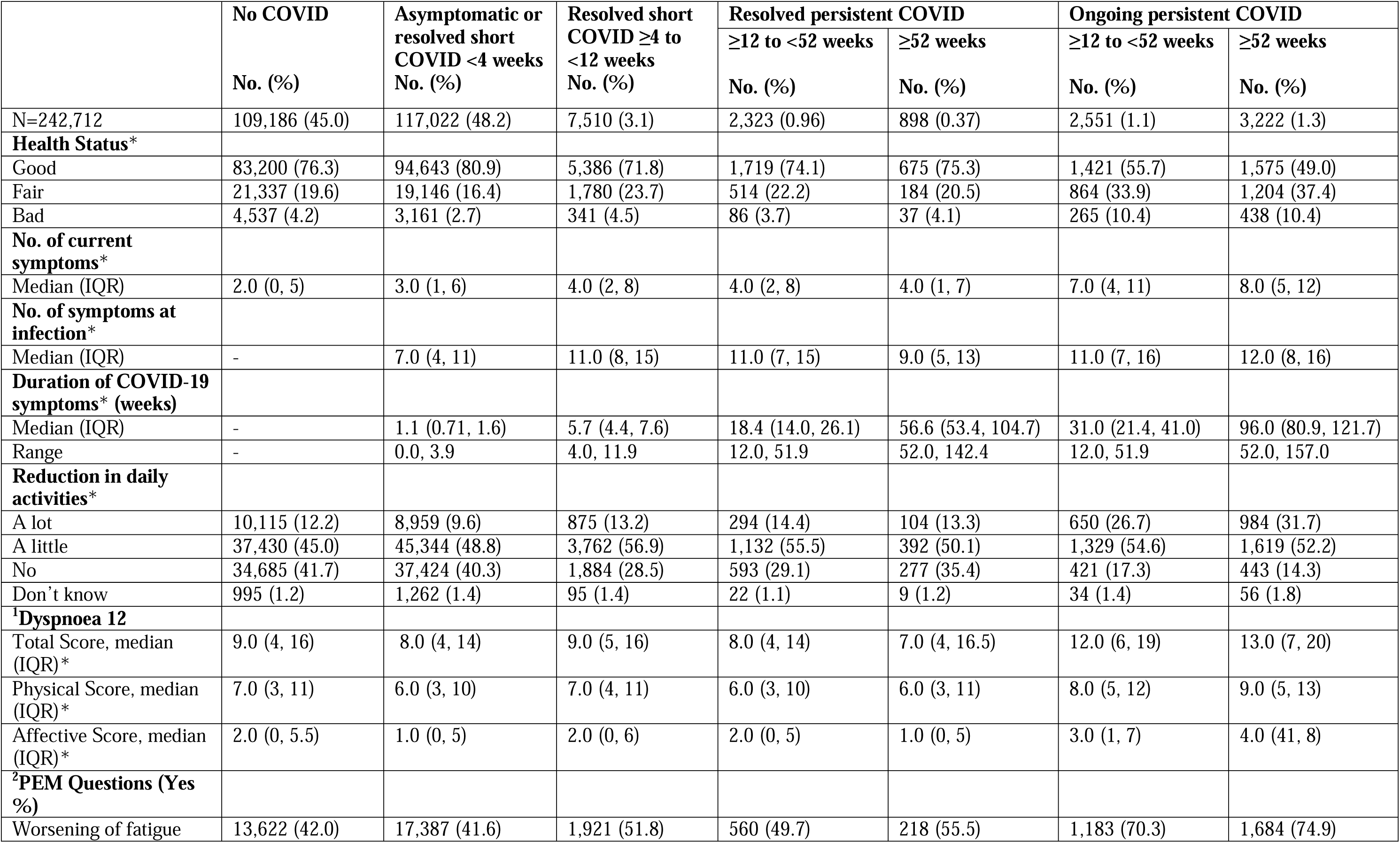
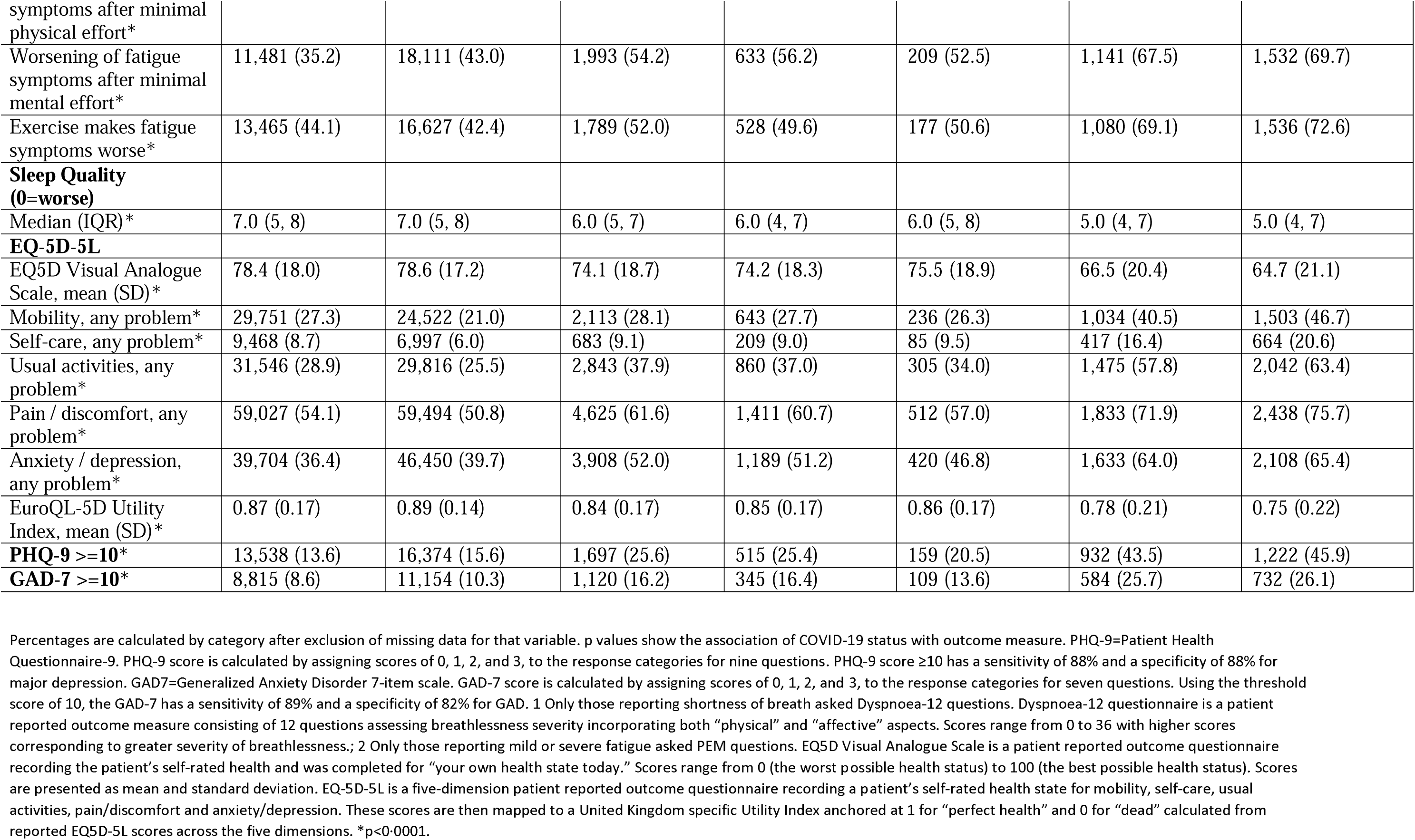
Current symptom profile and health-related quality of life of participants by COVID-19 history (n=242,712)

## DISCUSSION

This community-based study in England among adults aged ≥18 years describes patterns of symptoms and current health and well-being in a large cohort of people with a history of COVID-19 and compares individuals with ongoing COVID-19 symptoms for ≥12 weeks with those with no previous diagnosis of SARS-CoV-2 infection and those reporting resolution of COVID-19 symptoms.

### Duration of symptoms and risks of Long Covid

We show that symptomatic SARS-CoV-2 infection in England in adults is usually short-lived with most people reporting a short illness with symptom resolution within 2 weeks. However, one in 10 report symptoms for more than 4 weeks, one in 13 for more than 12 weeks (meeting the WHO definition for “post COVID-19 condition (Long COVID)” (22)), and 1 in 20 for more than 52 weeks. Our data suggest that 69% of those with persistent symptoms at 12 weeks will still have symptoms at 52 weeks, which we refer to as Very Long COVID. We found female sex, higher deprivation, having a pre-existing health condition, more severe symptoms at onset, and being infected when the original Wild-type variant was dominant to be associated with having COVID-19 symptoms persisting for ≥12 weeks and ≥52 weeks. The risk of persistent symptoms at 12 weeks was slightly lower in older than younger people; this is in contrast to other studies and may reflect the higher proportion of older people who are asymptomatic (4). The variant at the time of infection, initial severity and presence of pre-existing health conditions had the biggest impact on persistent symptoms, consistent with previous findings (4, 23, 24). Those infected when Omicron was dominant were 88% less likely to report symptoms beyond 12 weeks; this is likely to reflect changing immunity in the population from previous exposure to the virus and vaccination. A recent case-control study conducted in the UK found lower odds of Long COVID with the Omicron versus the Delta variant, ranging from OR 0.2 (95% CI 0.2–0·3) in those vaccinated >6 months prior to infection to 0.5 (95% CI 0.4–0.6) in those vaccinated <3 months prior to infection (25). We did not find conclusive evidence of effectiveness of vaccination against Long COVID. Vaccination reduces the severity of COVID-19 (26) and it may be through this indirect route that it impacts on the risk of persistent symptoms post-infection. A recent systematic review has suggested that vaccination before SARS-CoV-2 infection could reduce the risk of subsequent Long COVID (27).

### Common and specific symptoms

The reporting of symptoms was high across all groups in our study, including those who had never had COVID-19 and those who had recovered quickly. This may reflect the timing of the survey which included months with high levels of upper-respiratory and influenza-like illness in the population but emphasises the importance of having a comparison group to interpret reported symptom frequency in people with Long COVID. For example, while 66.9%, 54.9% and 54.6% of individuals reporting ongoing persistent COVID-19 symptoms reported currently experiencing mild fatigue, difficulty thinking or concentrating and joint pains, respectively, the prevalence of these symptoms in those with no history or evidence of previous SARS-CoV-2 infection was also high, at 31.1%, 15.2% and 35.5%. Indeed, this was also observed for those reporting they had recovered from COVID-19. These figures could be interpreted against pre-pandemic norms; however, we could not find any such published study in adults, and this would not take into account period changes in background symptomatology. These high rates compare to a much lower background prevalence of symptoms in PCR test negative participants (for example, mild fatigue 12.2%) who took part in the REACT-1 study rounds between August and December in 2020 and 2021 (see supplementary table S6). These findings of higher than expected symptom prevalence in comparison groups has been observed in other studies (28, 29) and could be due to self-selection by participants with symptoms to report, or returning to normal social mixing patterns following relaxation of social distancing measures in England from autumn 2021 (with exposure to other infections) or reflect the general physical and mental health impacts of living through the COVID-19 pandemic. However, our data did show that individuals with ongoing persistent COVID-19 symptoms were 9 times more likely to be experiencing loss or change of sense of smell and taste compared with other participants; shortness of breath, severe fatigue and difficulty thinking or concentrating were 7, 6 and 5 times more likely, indicating that these are more specific to Long COVID. Several systematic reviews, including one that only included community-based adults with mild acute COVID illness, support our findings by consistently reporting fatigue, shortness of breath, difficulty concentrating, anosmia and ageusia as some of the most common persistent COVID-19 symptoms (30–32). Of the few studies with a COVID-19-negative comparator group, one study showed that COVID-19 cases had a significantly higher likelihood of mood disorder, anxiety, and insomnia when compared to matched cohorts with influenza or respiratory tract infection (33). Another study found that COVID-19 cases had a significantly higher prevalence of symptoms at 6-and 9-months compared to community controls, including fatigue, sleep difficulties, hair loss, smell disorder, taste disorder, palpitations, chest pain, and headaches (34).

### Health and quality of life

There were substantial differences in currently reported health and well-being between individuals reporting ongoing persistent COVID-19 symptoms and those who had never had COVID-19 or had recovered, consistent with published evidence (23, 35). Encouragingly, those who had recovered from Long COVID, even after 52 weeks of symptoms, had scores for general health and quality of life similar to those with no COVID-19 or who recovered quickly. The dyspnoea and post-exertional malaise scales were asked of everyone reporting shortness of breath (Dyspnoea 12) or fatigue (PEM), and individuals reporting persistent COVID-19 symptoms scored higher than others, suggesting a specificity of these in Long COVID. A meta-analysis of 12 studies that evaluated health-related quality of life in individuals with Long COVID reported a pooled prevalence of poor quality of life (EQ5D Visual Analogue Scale - EQLJVAS) of 59% (95% CI 42, 75) (35). Based on individual factors in the EQLJ5DLJ5L questionnaire, the prevalence of problems with mobility was (36%; 95% CI 10, 67), self-care (8%; 95% CI 1, 21), usual activities (28%, 95% CI 2, 65), pain/discomfort (42%; 95% CI 28, 55), and anxiety/depression (38%, 95% CI 19, 58) (35).

In our study, persistent physical and mental health symptoms post-COVID-19 were shown to have an impact on quality of life and the ability to carry out day-to-day activities. This has implications for employment as chronically ill adults, irrespective of underlying disease, have considerably lower employment rates than healthy individuals (36). We did not collect data on working status, however the UK Office for National Statistics (ONS) have shown that working-age adults are less likely to participate in the labour market after developing Long COVID than they were before being infected with SARS-CoV-2 (37). Long COVID may therefore have contributed to the decreasing levels of participation seen in the UK labour market during the pandemic (37).

### Strengths and limitations

A strength of our study is its comparison of contemporaneous symptom profiles of community-based adults reporting ongoing persistent symptoms post-COVID-19 versus those who have never had COVID-19 or have recovered. Indeed, our findings highlight the importance of having a comparator cohort of participants who tested negative and experienced a pandemic and national lockdowns. However, we acknowledge the possibility of misclassification bias in our comparator groups as infections may have gone undetected particularly in the later stages of the pandemic when free universal testing was withdrawn by the UK Government. Also, symptom resolution post-COVID-19 was self-reported and therefore subjectiveness of recovery or symptom duration post-COVID-19 could also have led to misclassification. We used information regarding presence and duration of symptoms rather than whether participants described themselves as having “Long COVID” to reduce potential reporting bias. The data on symptoms at the time of PCR testing were retrospective which introduces the possibility of recall bias. However, previously we have shown that REACT participant reports of symptom onset date produce an epidemic curve that very closely tracks the epidemic (5). We used validated instruments to measure mental health (11, 12), quality of life (8), dyspnoea (9) and fatigue (38) but recognise the limitations of self-reporting and floor and ceiling effects (i.e, if a higher percentage of individuals achieve either maximum or minimum scores).

Our questionnaire response rate was 34.6%, higher than similar population-based COVID-19 epidemiological studies (1, 29, 37, 39). However, like these studies, our participants were more likely to be female, older, white ethnicity and from the least deprived areas compared with the general adult population. These issues might cause selection bias in our study; however, we did not observe substantial differences between those invited and those who participated in the study on the measured sociodemographic characteristics (see supplementary table 1).

Finally, public involvement from individuals with lived experience of Long COVID, including in generating item content for the questionnaire undoubtedly strengthened our design.

## Conclusion

Our study provides timely data about the effect and implications of the pandemic on adults in England with and without ongoing persistent COVID-19 symptoms. Although COVID-19 is usually of short duration, some adults experience persistent and burdensome illness. The multiple and varied symptoms imply that a multicomponent intervention will be needed to support and manage adults with Long COVID, building on existing management of other long-term conditions with a wide range of symptoms such as chronic fatigue syndrome. This is the first report from the REACT-Long COVID study, which includes bio-sampling for multi-omic, and biological endpoints. Further assessment of the trajectory of recovery of our participants, combined with this better mechanistic understanding, will inform a precision medicine approach to management of individuals with Long COVID.

## Data Availability

All data produced in the present study are available upon reasonable request to the authors.

