## Supplemental Tables & Figures for "LONG-TERM PHYSICAL AND MENTAL HEALTH IMPACT OF COVID-19 ON ADULTS IN ENGLAND: FOLLOW UP OF A LARGE RANDOM COMMUNITY SAMPLE"

**Figure S1: Study population flowchart**

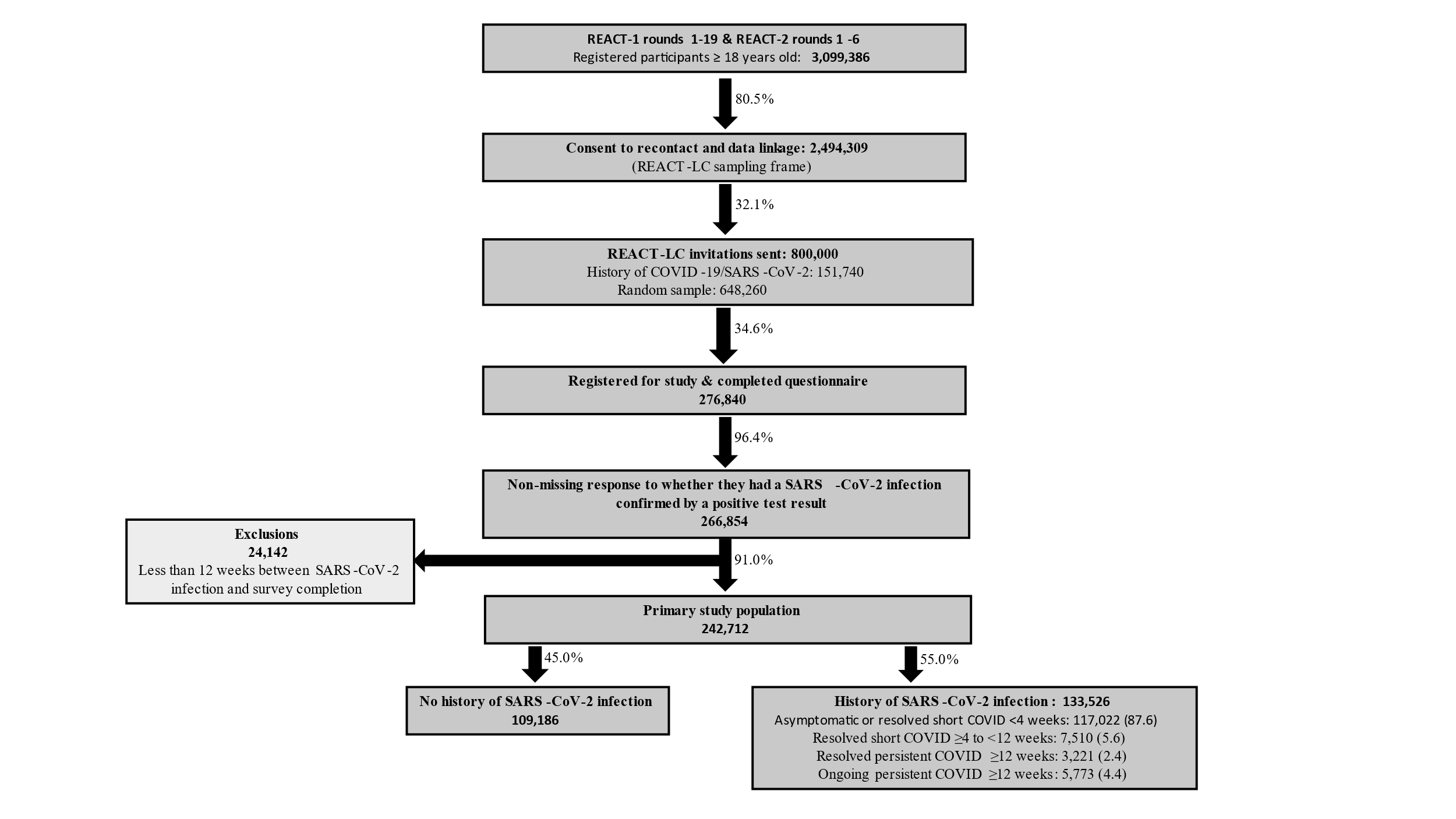

**Table S1: Sample frame and response of participants**

|  | **England (18+ population)^1, 2^** | **REACT-1/-2 (18+ population)** | **Target population** | **REACT-LC participants** |
| --- | --- | --- | --- | --- |
|  | N= 43,752,473 | N=3,099,386 | N=800,000 | N=276,840 |
| **Sex** |  |  |  |  |
| Male | 48.9 | 44.9 | 43.2 | 41.3 |
| Female | 51.1 | 55.1 | 56.8 | 58.7 |
| **Age** |  |  |  |  |
| 18-24 | 10.9 | 5.7 | 5.0 | 2.8 |
| 25-34 | 17.3 | 12.2 | 10.7 | 6.7 |
| 35-44 | 16.1 | 15.3 | 15.1 | 11.5 |
| 45-54 | 17.5 | 18.4 | 18.8 | 17.4 |
| 55-64 | 15.0 | 21.4 | 22.4 | 25.9 |
| 65-74 | 12.6 | 18.4 | 18.8 | 24.8 |
| 75+ | 10.5 | 8.5 | 9.3 | 10.9 |
| **Ethnicity** |  |  |  |  |
| White | 86.3 | 91.8 | 93.5 | 94.6 |
| Mixed | 1.1 | 1.3 | 1.3 | 1.2 |
| Asian | 7.3 | 4.5 | 3.5 | 2.7 |
| Black | 3.3 | 1.4 | 0.9 | 0.8 |
| Other | 1.9 | 0.93 | 0.8 | 0.7 |
| **Region** |  |  |  |  |
| North East | 4.8 | 3.8 | 3.9 | 3.8 |
| North West | 13.0 | 11.9 | 11.7 | 11.0 |
| Yorkshire and The Humber | 9.8 | 6.7 | 7.4 | 7.5 |
| East Midlands | 8.7 | 12.7 | 11.5 | 11.2 |
| West Midlands | 10.5 | 9.4 | 9.4 | 9.1 |
| East of England | 11.1 | 14.4 | 13.7 | 13.7 |
| London | 15.6 | 9.9 | 11.0 | 10.9 |
| South East | 16.3 | 21.7 | 21.3 | 21.9 |
| South West | 10.2 | 9.6 | 10.1 | 10.8 |
| **IMD Quintile** |  |  |  |  |
| 1 – most deprived | 19.8 | 11.2 | 9.8 | 8.4 |
| 2 | 20.9 | 16.8 | 15.8 | 15.1 |
| 3 | 20.4 | 21.5 | 21.5 | 21.4 |
| 4 | 19.7 | 24.2 | 24.9 | 25.4 |
| 5 – least deprived | 19.1 | 26.4 | 28.0 | 29.8 |

1 Office_for_National_Statistics. Population estimates for the UK, England and Wales, Scotland and Northern Ireland: mid2019 2020 [Available from: https://www.ons.gov.uk/releases/populationestimatesfortheukenglandandwalesscotlandandnorthernirelandmid2019]

2 Office_for_National_Statistics. Employee earnings in the UK: 2019 2019 [Available from: <https://www.ons.gov.uk/releases/employeeearningsintheuk2019>]

**Table S2: Comparison of participant sociodemographic and COVID-19 characteristics by COVID-19 history (n=** **242,712)**

|  | **N (%)** | **No COVID**  **No. (%)** | **Asymptomatic or resolved short COVID <4 weeks**  **No. (%)** | **Resolved short COVID ≥4 to <12 weeks**  **No. (%)** | **Resolved persistent COVID** | | **Ongoing persistent COVID** | |
| --- | --- | --- | --- | --- | --- | --- | --- | --- |
|  |  |  |  |  | **≥12 to <52 weeks**  **No. (%)** | **≥52 weeks**  **No. (%)** | **≥12 to <52 weeks**  **No. (%)** | **≥52weeks**  **No. (%)** |
|  | 242,712 | 109,186 (45.0) | 117,022 (48.2) | 7,510 (3.1) | 2,323 (0.96) | 898 (0.37) | 2,551 (1.1) | 3,222 (1.3) |
| **Age*** |  |  |  |  |  |  |  |  |
| 18 to 24 | 6,747 (2.8) | 2,311 | 3,887 | 199 | 90 | 43 | 84 | 133 |
|  |  | 34.3 | 57.6 | 3.0 | 1.33 | 0.64 | 1.24 | 1.97 |
| 25 to 34 | 16,040 (6.6) | 4,811 | 9,831 | 590 | 195 | 64 | 279 | 270 |
|  |  | 30.0 | 61.3 | 3.7 | 1.22 | 0.40 | 1.74 | 1.68 |
| 35 to 44 | 27,857 (11.5) | 7,965 | 16,971 | 1,272 | 402 | 140 | 536 | 571 |
|  |  | 28.6 | 60.9 | 4.6 | 1.44 | 0.50 | 1.92 | 2.05 |
| 45 to 54 | 42,264 (17.4) | 14,936 | 23,319 | 1,779 | 595 | 180 | 607 | 848 |
|  |  | 35.3 | 55.2 | 4.2 | 1.41 | 0.43 | 1.44 | 2.01 |
| 55 to 64 | 62,698 (25.8) | 28,181 | 30,139 | 2,023 | 636 | 249 | 585 | 885 |
|  |  | 45.0 | 48.1 | 3.2 | 1.01 | 0.40 | 0.93 | 1.41 |
| 65 to 74 | 60,411 (24.9) | 33,063 | 24,828 | 1,290 | 319 | 172 | 334 | 405 |
|  |  | 54.7 | 41.1 | 2.1 | 0.53 | 0.28 | 0.55 | 0.67 |
| 75+ | 26,695 (11.0) | 17,919 | 8,047 | 357 | 86 | 50 | 126 | 110 |
|  |  | 67.1 | 30.1 | 1.3 | 0.32 | 0.19 | 0.47 | 0.41 |
| **Sex at birth*** |  |  |  |  |  |  |  |  |
| Male | 100,898 (41.6) | 48,308 | 47,368 | 2,473 | 760 | 332 | 702 | 955 |
|  |  | 47.9 | 47.0 | 2.5 | 0.75 | 0.33 | 0.70 | 0.95 |
| Female | 141,807 (58.4) | 60,874 | 69,651 | 5,037 | 1,563 | 566 | 1,849 | 2,267 |
|  |  | 42.9 | 49.1 | 3.6 | 1.10 | 0.40 | 1.30 | 1.60 |
| **Ethnicity*** |  |  |  |  |  |  |  |  |
| White | 227,112 (94.6) | 102,265 | 109,600 | 6,895 | 2,137 | 829 | 2,387 | 2,999 |
|  |  | 45.0 | 48.3 | 3.0 | 0.94 | 0.37 | 1.05 | 1.32 |
| Mixed | 2,775 (1.2) | 1,007 | 1,507 | 118 | 47 | 10 | 45 | 41 |
|  |  | 36.3 | 54.3 | 4.3 | 1.69 | 0.36 | 1.62 | 1.48 |
| Asian | 6,435 (2.7) | 2,840 | 3,085 | 260 | 70 | 28 | 65 | 87 |
|  |  | 44.1 | 47.9 | 4.0 | 1.09 | 0.44 | 1.01 | 1.35 |
| Black | 1,987 (0.83) | 985 | 846 | 79 | 22 | 12 | 14 | 29 |
|  |  | 49.6 | 42.6 | 4.0 | 1.11 | 0.60 | 0.70 | 1.46 |
| Other | 1,697 (0.71) | 692 | 833 | 87 | 30 | 9 | 20 | 26 |
|  |  | 40.8 | 49.1 | 5.1 | 1.77 | 0.53 | 1.18 | 1.53 |
| **IMD*** |  |  |  |  |  |  |  |  |
| 1 – most deprived | 19,888 (8.4) | 8,604 | 9,473 | 744 | 227 | 95 | 276 | 469 |
|  |  | 43.3 | 47.6 | 3.7 | 1.14 | 0.48 | 1.39 | 2.36 |
| 2 | 35,564 (15.0) | 15,847 | 17,063 | 1,146 | 369 | 130 | 429 | 580 |
|  |  | 44.6 | 48.0 | 3.2 | 1.04 | 0.37 | 1.21 | 1.63 |
| 3 | 50,627 (21.4) | 23,183 | 24,032 | 1,547 | 500 | 196 | 509 | 660 |
|  |  | 45.8 | 47.5 | 3.1 | 0.99 | 0.39 | 1.01 | 1.30 |
| 4 | 60,165 (25.4) | 27,187 | 29,103 | 1,794 | 531 | 197 | 617 | 736 |
|  |  | 45.2 | 48.4 | 3.0 | 0.88 | 0.33 | 1.03 | 1.22 |
| 5 – least deprived | 70,419 (29.8) | 31,722 | 34,294 | 2,115 | 652 | 254 | 668 | 714 |
|  |  | 45.1 | 48.7 | 3.0 | 0.93 | 0.36 | 0.95 | 1.01 |
| **Comorbidities*** |  |  |  |  |  |  |  |  |
| 0 | 156,433 (64.5) | 34,593 | 106,790 | 6,920 | 2,153 | 843 | 2,254 | 2,880 |
|  |  | 22.1 | 68.3 | 4.4 | 1.38 | 0.54 | 1.44 | 1.84 |
| 1 | 38,811 (16.0) | 30,347 | 7,402 | 425 | 143 | 41 | 220 | 233 |
|  |  | 78.2 | 19.1 | 1.1 | 0.37 | 0.11 | 0.57 | 0.60 |
| 2 or more | 47,468 (19.6) | 44,246 | 2,830 | 165 | 27 | 14 | 77 | 109 |
|  |  | 93.2 | 6.0 | 0.35 | 0.06 | 0.03 | 0.16 | 0.23 |
| **Smoking Status*** |  |  |  |  |  |  |  |  |
| Yes | 13,257 (5.7) | 7,096 | 5,365 | 333 | 85 | 31 | 165 | 182 |
|  |  | 53.5 | 40.5 | 2.5 | 0.64 | 0.23 | 1.24 | 1.37 |
| No | 217,796 (94.3) | 98,104 | 105,037 | 6,795 | 2,080 | 785 | 2,200 | 2,795 |
|  |  | 45.0 | 48.2 | 3.1 | 0.96 | 0.36 | 1.01 | 1.28 |
| **Severity of initial SARS-CoV-2 infection*** |  |  |  |  |  |  |  |  |
| No symptoms | 6,206 (4.7) | - | 6,206 | 0 | 0 | 0 | 0 | 0 |
|  |  |  | 100.0 | 0.0 | 0.00 | 0.00 | 0.00 | 0.00 |
| Mild symptoms | 36,207 (27.2) | - | 34,346 | 724 | 228 | 230 | 345 | 334 |
|  |  |  | 94.9 | 2.00 | 0.63 | 0.64 | 0.95 | 0.92 |
| Moderate symptoms | 64,958 (48.8) | - | 58,023 | 3,308 | 916 | 404 | 1,174 | 1,133 |
|  |  |  | 89.3 | 5.1 | 1.41 | 0.62 | 1.81 | 1.74 |
| Severe symptoms | 25,682 (19.3) | - | 17,974 | 3,478 | 1,179 | 264 | 1,032 | 1,755 |
|  |  |  | 70.0 | 13.5 | 4.59 | 1.03 | 4.02 | 6.83 |
| **Dominant strain at symptom onset or positive test*** |  |  |  |  |  |  |  |  |
| Wild type (before Dec 2020) | 15,174 (11.6) | - | 10,157 | 1,924 | 908 | 542 | 0 | 1,643 |
|  |  |  | 66.9 | 12.7 | 5.98 | 3.57 | 0.00 | 10.83 |
| Alpha (Dec 2020-April 2021) | 12,054 (9.2) | - | 9,124 | 1,264 | 508 | 260 | 0 | 898 |
|  |  |  | 75.7 | 10.5 | 4.21 | 2.16 | 0.00 | 7.45 |
| Delta (May 2021-mid-Dec 2021) | 19,993 (15.3) | - | 16,679 | 1,464 | 462 | 96 | 611 | 681 |
|  |  |  | 83.4 | 7.3 | 2.31 | 0.48 | 3.06 | 3.41 |
| Omicron (after mid-Dec 2021) | 83,544 (63.9) | - | 78,301 | 2,858 | 445 | 0 | 1,940 | 0 |
|  |  |  | *93.7* | 3.4 | 0.53 | 0.00 | 2.32 | 0.00 |
| **Vaccination status at symptom onset or positive test*** |  |  |  |  |  |  |  |  |
| 0 | 67,013 (50.2) | - | 56,079 | 4,653 | 1,759 | 812 | 844 | 2,866 |
|  |  |  | 83.7 | 6.9 | 2.62 | 1.21 | 1.26 | 4.28 |
| 1 | 2,147 (1.6) | - | 1,804 | 125 | 44 | 39 | 24 | 111 |
|  |  |  | 84.0 | 5.8 | 2.05 | 1.82 | 1.12 | 5.17 |
| 2 or more | 64,366 (48.2) | - | 59,139 | 2,732 | 520 | 47 | 1,683 | 245 |
|  |  |  | 91.9 | 4.2 | 0.81 | 0.07 | 2.61 | 0.38 |

Percentages are calculated by category after exclusion of missing data for that variable; p values show the association of COVID-19 status with sociodemographic or COVID-19 characteristic. *p<0·0001

**Table S3: Factors associated with persistent COVID-19 symptoms lasting i) ≥12 weeks and ii) ≥52 weeks.**

Odds ratio compares participants with persistent symptoms lasting i) ≥12 weeks or ii) ≥52 weeks with those who reported being asymptomatic or symptoms resolved within 4 weeks.

|  | **Controls:**  **asymptomatic or resolved short COVID <4 weeks** | **Cases 1: persistent COVID ≥12 weeks** | **Unadjusted OR**  **(95% CI)** | **^$^Adjusted OR**  **(95% CI)** | **Cases 2: persistent COVID ≥52 weeks** | **Unadjusted OR**  **(95% CI)** | **^$^Adjusted OR**  **(95% CI)** |
| --- | --- | --- | --- | --- | --- | --- | --- |
|  | N=117,022 | N=8,994 |  |  | N= 4,120 |  |  |
| **Age** |  |  |  |  |  |  |  |
| 18 to 24 | 3,887 | 350 | REF | REF | 176 | REF | REF |
|  | 3.32 | 3.89 |  |  | 4.27 |  |  |
| 25 to 34 | 9,831 | 808 | 0.91 (0.80, 1.04) | 0.95 (0.82, 1.11) | 334 | 0.75 (0.62, 0.90)** | 0.86 (0.69, 1.07) |
|  | 8.40 | 8.98 |  |  | 8.11 |  |  |
| 35 to 44 | 16,971 | 1,649 | 1.08 (0.96, 1.22) | 1.12 (0.97, 1.28) | 711 | 0.93 (0.78, 1.10) | 1.10 (0.90, 1.34) |
|  | 14.50 | 18.33 |  |  | 17.26 |  |  |
| 45 to 54 | 23,319 | 2,230 | 1.06 (0.94, 1.19) | 1.00 (0.88, 1.15) | 1,028 | 0..97 (0.83, 1.15) | 1.02 (0.84, 1.23) |
|  | 19.93 | 24.79 |  |  | 24.95 |  |  |
| 55 to 64 | 30,139 | 2,355 | 0.87 (0 .77, 0.98)* | 0.87 (0.75, 0.99)* | 1,134 | 0.83 (0.71, 0.98)* | 0.92 (0.75, 1.11) |
|  | 25.75 | 26.18 |  |  | 27.52 |  |  |
| 65 to 74 | 24,828 | 1,230 | 0.55 (0.49, 0.62)*** | 0.74 (0.64, 0.86)*** | 577 | 0.51 (0.43, 0.61)*** | 0.84 (0.69, 1.03) |
|  | 21.22 | 13.68 |  |  | 14.00 |  |  |
| 75+ | 8,047 | 372 | 0.51 (0.44, 0.60)*** | 0.84 (0.70, 0.99)* | 160 | 0.44 (0.35, 0.55)*** | 0.86 (0.66, 1.11) |
|  | 6.88 | 4.14 |  |  | 3.88 |  |  |
| **Sex at birth** |  |  |  |  |  |  |  |
| Male | 47,368 | 2,749 | REF | REF | 1,287 | REF | REF |
|  | 40.48 | 30.56 |  |  | 31.24 |  |  |
| Female | 69,651 | 6,245 | 1.54 (1.47, 1.62)*** | 1.42 (1.35, 1.50)*** | 2,833 | 1.50 (1.40, 1.60)*** | 1.49 (1.38, 1.62)*** |
|  | 59.52 | 69.44 |  |  | 68.76 |  |  |
| **Ethnicity** |  |  |  |  |  |  |  |
| White | 109,600 | 8,352 | REF | REF | 3,828 | REF | REF |
|  | 94.59 | 93.77 |  |  | 94.05 |  |  |
| Mixed | 1,507 | 143 | 1.25 (1.04, 1.48)* | 0.99 (0.81, 1.21) | 51 | 0.97 (0.73, 1.28) | 0.71 (0.51, 1.00) |
|  | 1.30 | 1.61 |  |  | 1.25 |  |  |
| Asian | 3,085 | 250 | 1.06 (0.93, 1.21) | 0.80 (0.69 , 0.93)** | 115 | 1.07 (0.88, 1.29) | 0.71 (0.57, 0.88)** |
|  | 2.66 | 2.81 |  |  | 2.83 |  |  |
| Black | 846 | 77 | 1.19 (0.94, 1.51) | 0.93 (0.71, 1.21) | 41 | 1.39 (1.01, 1.90)* | 0.95 (0.66, 1.37) |
|  | 0.73 | 0.86 |  |  | 1.01 |  |  |
| Other | 833 | 85 | 1.34 (1.07, 1.68)* | 0.91 (0.70, 1.19) | 35 | 1.20 (0.86, 1.69) | 0.72 (0.48, 1.06) |
|  | 0.72 | 0.95 |  |  | 0.86 |  |  |
| **IMD** |  |  |  |  |  |  |  |
| 1: most deprived | 9,473 | 1,067 | REF | REF | 564 | REF | REF |
|  | 8.31 | 12.11 |  |  | 13.99 |  |  |
| 2 | 17,063 | 1,508 | 0.78 (0.72, 0.85)*** | 0.90 (0.82, 0.99)* | 710 | 0.70 (0.62, 0.78)*** | 0.87 (0.76, 0.99)* |
|  | 14.97 | 17.12 |  |  | 17.61 |  |  |
| 3 | 24,032 | 1,865 | 0.69 (0.64, 0.75)*** | 0.87 (0.79, 0.95)** | 856 | 0.60 (0.54, 0.67)*** | 0.83 (0.73, 0.95)** |
|  | 21.09 | 21.17 |  |  | 21.24 |  |  |
| 4 | 29,103 | 2,081 | 0.63 (0.59, 0.69)*** | 0.83 (0 76, 0.91)*** | 933 | 0.54 (0.48, 0.60)*** | 0.78 (0.69, 0.89)*** |
|  | 25.54 | 23.62 |  |  | 23.15 |  |  |
| 5: least deprived | 34,294 | 2,288 | 0.59 (0.55, 0.64)*** | 0 82 (0.75, 0.90)*** | 968 | 0.47 (0.43, 0.53)*** | 0.75 (0.66 , 0.85)*** |
|  | 30.09 | 25.97 |  |  | 24.01 |  |  |
| **Comorbidities** |  |  |  |  |  |  |  |
| 0 | 106,790 | 8,130 | REF | REF | 3,723 | REF | REF |
|  | 91.26 | 90.39 |  |  | 90.36 |  |  |
| 1 | 7,402 | 637 | 1.13 (1.04, 1.23) | 1.31 (1.19, 1.44)*** | 274 | 1.06 (0.94, 1.20) | 1.52 (1.31, 1.76)*** |
|  | 6.33 | 7.08 |  |  | 6.65 |  |  |
| 2 or more | 2,830 | 227 | 1.05 (0.92, 1.21) | 1.46 (1.27, 1.75)*** | 123 | 1.25 (1.04, 1.50)* | 2.35 (1.85, 2.97)*** |
|  | 2.42 | 2.52 |  |  | 2.99 |  |  |
| **Smoking Status** |  |  |  |  |  |  |  |
| Yes | 5,365 | 463 | REF | REF | 213 | REF | REF |
|  | 4.86 | 5.56 |  |  | 5.62 |  |  |
| No | 105,037 | 7,860 | 0.87 (0.79, 0.96)** | 0.98 (0.88, 1.09) | 3,580 | 0.86 (0.75, 0.99)* | 0.99 (0.84, 1.15) |
|  | 95.14 | 94.44 |  |  | 94.38 |  |  |
| **Severity of initial SARS-CoV-2 infection** |  |  |  |  |  |  |  |
| No symptoms | 6,206 | 0 | omitted | omitted | 0 | omitted | omitted |
|  | 5.32 | 0.0 |  |  | 0.00 |  |  |
| Mild symptoms | 34,346 | 1,137 | REF | REF | 564 | REF | REF |
|  | 29.47 | 12.64 |  |  | 13.69 |  |  |
| Moderate symptoms | 58,023 | 3,627 | 1.89 (1.76, 2.02)*** | 1.76 (1.63, 1.89)*** | 1,537 | 1.61 (1.46, 1.78)*** | 1.47 (1.32, 1.64)*** |
|  | 49.78 | 40.33 |  |  | 37.31 |  |  |
| Severe symptoms | 17,974 | 4,230 | 7.11 (6.64, 7.61)*** | 4.87 (4.52, 5.25)*** | 2,019 | 6.84 (6.22, 7.52)*** | 3.55 (3.19, 3.96)*** |
|  | 15.42 | 47.03 |  |  | 49.00 |  |  |
| **Dominant strain at symptom onset or positive test** |  |  |  |  |  |  |  |
| Wild type (before Dec 2020) | 10,157 | 3,093 | REF | REF | 2,185 | REF | REF |
|  | 8.89 | 34.39 |  |  | 53.03 |  |  |
| Alpha (Dec 2020-April 2021) | 9,124 | 1,666 | 0.60 (0.56, 0.64)*** | 0.60 (0.56, 0.64)*** | 1,158 | 0.59 (0.55, 0.64)*** | 0.59 (0.54, 0.64)*** |
|  | 7.99 | 18.52 |  |  | 28.11 |  |  |
| Delta (May 2021-mid-Dec 2021) | 16,679 | 1,850 | 0.36 (0.34, 0.39)*** | 0.38 (0.35, 0.41)*** | 777 | 0.22 (0.20, 0.24)*** | 0.32 (0.29, 0.36)*** |
|  | 14.60 | 20.57 |  |  | 18.86 |  |  |
| Omicron (after mid-Dec 2021) | 78,301 | 2,385 | 0.10 (0.09, 0.11)*** | 0.12 (0.11, 0.13)*** | 0 | Omitted | Omitted |
|  | 68.53 | 26.52 |  |  | 0.00 |  |  |
| **Vaccination status at symptom onset or positive test** |  |  |  |  |  |  |  |
| 0 | 56,079 | 6,281 | REF | REF | 3,678 | REF | REF |
|  | 47.92 | 69.84 |  |  | 89.27 |  |  |
| 1 | 1,804 | 218 | 1.08 (0.94, 1.24) | 0.98 (0.84, 1.15) | 150 | 1.27 (1.07, 1.50)** | 1.06 (0.88, 1.29) |
|  | 1.54 | 2.42 |  |  | 3.64 |  |  |
| 2 or more | 59,139 | 2,495 | 0.38 (0.36, 0.40)*** | 0.94 (0.87, 1.00) | 292 | 0.08 (0.07, 0.09)*** | 0.43 (0.37, 0.51)*** |
|  | 50.54 | 27.74 |  |  | 7.09 |  |  |

**^$^**Mutually adjusted for age, sex, ethnicity, IMD, comorbidities, smoking status, severity of initial infection, dominant variant at time of infection, vaccination status

**Table S4: Survival analysis with Accelerated Time Failure model: Factors associated with likelihood (adjusted Time Ratio) of longer duration of symptoms in study participants with symptomatic SARS-CoV-2 infection lasting ≥12 weeks (N=8,532)**

|  | N | Unadjusted TR  (95% CI) | ^$^Adjusted TR  (95% CI) |
| --- | --- | --- | --- |
| **Age** |  |  |  |
| 18 to 24 | 329 | REF | REF |
| 25 to 34 | 757 | 1.04 (0.85, 1.28) | 0.95 (0.77, 1.18) |
| 35 to 44 | 1,551 | 1.04 (0.86, 1.26) | 0.99 (0.81, 1.21) |
| 45 to 54 | 2,123 | 1.01 (0.84, 1.22) | 0.95 (0.78, 1.15) |
| 55 to 64 | 2,225 | 0.95 (0.79, 1.14) | 0.91 (0.75, 1.11) |
| 65 to 74 | 1,191 | 0.91 (0.75, 1.10) | 0.91 (0.74, 1.11) |
| 75+ | 356 | 0.97 (0.76, 1.23) | 0.98 (0.76, 1.26) |
| **Sex at birth** |  |  |  |
| Male | 2,613 | REF | REF |
| Female | 5,919 | 1.13 (1.05, 1.21)** | 1.14 (1.06, 1.23)** |
| **Ethnicity** |  |  |  |
| White | 7,917 | REF | REF |
| Mixed | 136 | 0.80 (0.62, 1.04) | 0.75 (0.57, 0.99)* |
| Asian | 242 | 0.87 (0.72, 1.06) | 0.85 (0.69, 1.05) |
| Black | 75 | 0.90 (0.64, 1.27) | 0.80 (0.56, 1.14) |
| Other | 78 | 0.69 (0.50, 0.96)** | 0.63 (0.45, 0.89)** |
| **IMD** |  |  |  |
| 1 – most deprived | 1,007 | REF | REF |
| 2 | 1,424 | 0.86 (0.76, 0.98)* | 0.91 (0.80, 1.04) |
| 3 | 1,768 | 0.79 (0.70, 0.89)*** | 0.81 (0.71, 0.92)** |
| 4 | 1,962 | 0.82 (0.72, 0.93)** | 0.89 (0.78, 1.01) |
| 5 – least deprived | 2,197 | 0.72 (0.64, 0.81)*** | 0.78 (0.69, 0.88)*** |
| **Comorbidities** |  |  |  |
| 0 | 7,704 | REF | REF |
| 1 | 612 | 1.21 (1.06, 1.39)** | 1.24 (1.08, 1.42)** |
| 2 or more | 216 | 2.11 (1.64, 2.71)*** | 2.05 (1.58, 2.66)*** |
| **Smoking Status** |  |  |  |
| Yes | 441 | REF | REF |
| No | 7,452 | 0.70 (0.59, 0.83)*** | 0.73 (0.62, 0.86)*** |
| **Severity of initial SARS-CoV-2 infection** |  |  |  |
| No symptoms | 0 | omitted | omitted |
| Mild symptoms | 1,016 | REF | REF |
| Moderate symptoms | 3,410 | 0.93 (0.84, 1.04) | 0.90 (0.81, 1.01) |
| Severe symptoms | 4,106 | 1.05 (0.94, 1.17) | 1.00 (0.90, 1.12) |
| **Dominant strain at symptom onset or positive test** |  |  |  |
| Wild type (before Dec 2020) | 3,074 | REF | REF |
| Alpha (Dec 2020-April 2021) | 1,635 | 0.82 (0.75, 0.90)*** | 0.79 (0.72, 0.86)*** |
| Delta (May 2021-mid-Dec 2021) | 1,786 | 0.89 (0.81, 0.97)* | 0.89 (0.79, 0.99)* |
| Omicron (after mid-Dec 2021) | 2,037 | 0.70 (0.63, 0.76)*** | 0.69 (0.61, 0.78)*** |
| **Vaccination status at symptom onset or positive test** |  |  |  |
| 0 | 6,068 | REF | REF |
| 1 | 208 | 1.00 (0.80, 1.23) | 1.10 (0.87, 1.38) |
| 2 or more | 2,256 | 0.86 (0.79, 0.93)*** | 0.98 (0.88, 1.10) |

*p<0·05, **p<0·01, ***p<0·0001

^$^ Mutually adjusted for age, sex, ethnicity, IMD, comorbidities, smoking status, severity of initial infection, dominant variant at time of infection, vaccination status.

**Table S5: Association between COVID-19 history and current symptom profile and health-related quality of life characteristics adjusted for age, sex, ethnicity, IMD, comorbidities, smoking status. Odds Ratios with 95% CI (White Cells) and Regression Coefficients, 95% CI (Grey Cells)**

|  | **No COVID** | **Asymptomatic or resolved short COVID <4 weeks**  **(Reference Group)** | **Resolved short COVID ≥4 to <12 weeks** | **Resolved persistent COVID ≥12 weeks** | **Ongoing persistent COVID ≥12 weeks** |
| --- | --- | --- | --- | --- | --- |
| **Health Status** |  |  |  |  |  |
| Good/Fair | - | - | - | - | - |
| Bad | 0.37 (0.34, 0.39) | - | 1.73 (1.53, 1.96) | 1.40 (1.13, 1.72) | 4.95 (4.50, 5.45) |
| **No. of current symptoms** | -1.72 (-1.76, -1.68) | - | 1.33 (1.25, 1.42) | 1.18 (1.06, 1.31) | 4.12 (4.02, 4.21) |
| **Reduction in daily activities** |  |  |  |  |  |
| No | - | - | - | - | - |
| Yes | 0.57 (0.55, 0.59) | - | 1.68 (1.58, 1.78) | 1.48 (1.36, 1.61) | 3.56 (3.29, 3.86) |
| **^1^Dyspnoea 12** |  |  |  |  |  |
| Total Score | -2.37 (-2.78, -1.96) | - | 1.16 (0.58, 1.74) | 0.16 (-0.71, 1.03) | 4.03 (3.57, 4.49) |
| Physical Score | -1.29 (-1.53, -1.05) | - | 0.67 (0.34, 1.01) | 0.23 (-0.28, 0.74) | 2.20 (1.93, 2.47) |
| Affective Score | -1.02 (-1.20, -0.84) | - | 0.47 (0.21, 0.72) | 0.05 (-0.34, 0 .44) | 1.82 (1.62, 2.02) |
| **^2^PEM Questions** |  |  |  |  |  |
| Worsening of fatigue symptoms after minimal physical effort |  |  |  |  |  |
| No | - | - | - | - | - |
| Yes | 0.46 (0.44, 0.49) | - | 1.54 (1.43, 1.65) | 1.48 (1.33, 1.66) | 3.87 (3.58, 4.18) |
| Worsening of fatigue symptoms after minimal mental effort |  |  |  |  |  |
| No | - | - | - | - | - |
| Yes | 0.53 (0.51, 0.56) | - | 1.61 (1.49, 1.73) | 1.57 (1.41, 1.76) | 2.92 (2.70, 3.15) |
| Exercise makes fatigue symptoms worse |  |  |  |  |  |
| No | - | - | - | - | - |
| Yes | 0.53 (0.50 , 0.56) | - | 1.49 (1.39, 1.61) | 1.34 (1.20, 1.50) | 3.44 (3.18, 3.72) |
| **Sleep Quality** | 0.52 (0.50, 0.54) | - | -0.38 (-0.43, -0.34) | -0.46 (-0.53, -0.39) | -0.91 (-0.96, -0.86) |
| **EQ-5D-5L** |  |  |  |  |  |
| EQ5D Visual Analogue | 6.24 (6.05, 6.42) | - | -4.35 (-4.75, -3.94) | -3.75 (-4.37, -3.14) | -12.23 (-12.70, -11.77) |
| Mobility: Any problem |  |  |  |  |  |
| No | - | - | - | - | - |
| Yes | 0.45 (0.44, 0.47) | - | 1.58 (1.49, 1.67) | 1.53 (1.40, 1.67) | 3.39 (3.19, 3.59) |
| Self-care: Any problem |  |  |  |  |  |
| No | - | - | - | - | - |
| Yes | 0.43 (0.41, 0.45) | - | 1.61 (1.48, 1.76) | 1.68 (1.47, 1.92) | 3.63 (3.36, 3.93) |
| Usual activities: Any problem |  |  |  |  |  |
| No | - | - | - | - | - |
| Yes | 0.40 (0.39, 0.41) | - | 1.84 (1.75, 1.94) | 1.74 (1.61, 1.88) | 4.86 (4.58, 5.15) |
| Pain / discomfort: Any problem |  |  |  |  |  |
| No | - | - | - | - | - |
| Yes | 0.51 (0.49, 0.52) | - | 1.58 (1.50, 1.66) | 1.49 (1.38, 1.61) | 2.90 (2.72, 3.09) |
| Anxiety / depression: Any problem |  |  |  |  |  |
| No | - | - | - | - | - |
| Yes | 0.42 (0.41, 0.44) | - | 1.64 (1.56,1.72) | 1.44 (1.34, 1.56) | 2.53 (2.38, 2.69) |
| EuroQL-5D Utility Index | 0.06 (0.054, 0.057) | - | -0.04 (-0.044, -0.037) | -0.03 (-0.040, -0.029) | -0.12 (-.121, -0.113) |
| **PHQ-9 (>=10)** |  |  |  |  |  |
| No | - | - | - | - | - |
| Yes | 0.35 (0.34, 0.37) | - | 1.88 (1.76, 2.00) | 1.70 (1.54, 1.86) | 4.11 (3.85, 4.39) |
| **GAD-7 (>=10)** |  |  |  |  |  |
| No | - | - | - | - | - |
| Yes | 0.36 (0.34, 0.37) | - | 1.68 (1.56, 1.80) | 1.56 (1.40, 1.74) | 2.72 (2.53, 2.93) |

**Table S6: Prevalence of symptoms persisting for 11 days or more in 123,468 PCR negative adults aged ≥18 years in REACT-1**

To estimate background prevalence of symptoms, we used data on history of symptoms lasting 11 or more days (that were present within the 7 days prior to questionnaire completion) for individuals aged ≥18 years who had a negative PCR test (N=831,424) in the REACT-1 study. The data were weighted (by sex, age, ethnicity, Lower Tier Local Authority population and IMD) to take account of the sampling design and differential response rates, to obtain prevalence estimates that were representative of the adult population of England. Data collected between August and December in 2020 and 2021 (rounds 5-7 and-14-16).

| **Symptom** | ***% (95% CI)*** |
| --- | --- |
| Loss or change of sense of smell | 0.88 (0.85, 0.91) |
| Loss or change of sense of taste | 0.69 (0.66, 0.71) |
| Fever | 2.6 (2.5, 2.7) |
| Coughing | 3.2 (3.1, 3.3) |
| Itchy, sore or red eyes, conjunctivitis | 3.8 (3.7, 3.9) |
| Leg swelling (Thrombosis) | 0.38 (0.36, 0.42) |
| Appetite loss | 2.5 (2.4, 2.6) |
| Dizziness, vertigo | 4.0 (3.9, 4.1) |
| Headaches | 12.8 (12.7, 12.9) |
| Shortness of breath, breathlessness, wheezing | 3.9 (3.8, 4.0) |
| Tightness or heaviness in chest, chest pain | 3.6 (3.5, 3.7) |
| Difficulty sleeping | 7.3 (7.2, 7.4) |
| Mild fatigue (e.g. feeling tired) | 12.2 (12.1, 12.3) |
| Severe fatigue (e.g. inability to get out of bed) | 1.5 (1.4, 1.6) |
| Numbness or tingling somewhere in the body | 2.6 (2.5, 2.7) |
| Achy or cramping muscles, pain in muscles | 7.0 (6.9, 7.1) |
